# Caries incidence in school-based oral health programs

**DOI:** 10.1101/2024.07.29.24311175

**Authors:** Ryan Richard Ruff

## Abstract

**Background:** School-based caries prevention can increase access to critical dental services and reduce oral health inequities. However, little is known regarding the incidence of dental caries in children participating in school caries prevention, and caries diagnosis is often interval censored.

**Methods:** In this paper, we used data from a longitudinal, school-based, randomized clinical trial of minimally invasive treatments for dental caries to estimate the per-visit incidence rate and compare the hazard of dental caries in children receiving either silver diamine fluoride or glass ionomer dental sealants. To account for interval censoring, we used semiparametric transformation models for univariate failure time data and imputed the caries incidence using G-imputation.

**Results:** There were 3040 children that met inclusion criteria for analysis, 1516 (49.9%) of which were randomly assigned to receive silver diamine fluoride and 1524 (50.1%) assigned to receive glass ionomer dental sealants. There were no differences in the hazard of caries between treatments (HR = 0.98, 95% CI = 0.73, 1.24), while children with caries at baseline had a significant increase in the hazard of new caries (HR = 2.54, 95% CI = 2.26, 2.83) compared to those that were caries-free. The per-visit caries incidence ranged from 4.8 to 11.1 per 1000 person-years and increased with each successive study observation.

**Conclusions:** School-based caries prevention can positively affect caries incidence, and results can be used to inform future program design and implementation.

## Introduction

Dental caries is the most prevalent noncommunicable disease in the world, and is highly inequitable across socioeconomic strata [1]. Children with untreated dental caries face considerable health and psychosocial sequela, including negative effects on quality of life [2] and educational performance [3]. If left untreated, caries can result in pain, chronic systemic infection [4], or adverse growth patterns [5]. The multifactorial etiology of dental caries, including behavioral, socioeconomic, physiological, and genetic contributions, poses a number of unique challenges for disease prevention [6].

A number of dental procedures can be provided within schools to prevent and manage the burden of caries, directly addressing a key determinant of disease. For example, the US Community Preventive Services Task Force officially recommends school-based sealant programs [7, 8], while other approaches can range from simple preventive care including examinations, cleanings, and fluoride treatments to more comprehensive services including x-rays, atraumatic restorations, and fillings [9]. More recently, silver diamine fluoride has been shown to be effective when used in school caries prevention [10]. Irrespective of the treatment provided, school-based caries prevention increases access to critical dental care for traditionally underserved groups and reduces oral health inequities.

By nature of their design, school prevention programs provide care at predetermined intervals, typically annually or biannually [11]. Due to this periodic follow-up, the actual time for caries onset is interval censored and all that can be derived is the time of the latest known negative diagnosis and the earliest known positive diagnosis. Interval censoring is a common problem in clinical trials and longitudinal research [12]. Subsequently, valid estimation of caries incidence in the presence of interval censoring can help identify optimal times for follow-up screening and treatment in school-based caries prevention.

The *CariedAway* study was a school-based randomized trial of minimally invasive treatments to prevent and control dental caries [13]. Originally designed to provide biannual care, the CariedAway implementation period coincided with the COVID-19 pandemic, resulting in both inconsistent care delivery to program participants and considerable interval censoring. Our objective was to estimate the per-visit caries incidence rate in a sample of high-risk, low-access children.

## Methods

### Design and Participants

CariedAway was a pragmatic, cluster-randomized, longitudinal non-inferiority trial. The original objectives of CariedAway were to compare the efficacy of silver diamine fluoride to dental sealants and atraumatic restorations in the two-year arrest and four-year prevention of caries. The study is registered at www.clinicaltrials.gov (#NCT03442309) and received ethical approval from the New York University School of Medicine Institutional Review Board. CariedAway was conducted from February 1, 2018 through June 1, 2023.

Any primary school that was part of the New York City public school system with a student population consisting of at least 50% Black and/or Hispanic/Latino and 80% receiving free and reduced lunch (as a proxy for low-income families) was eligible for participation. After school enrollment, all children were eligible for care if they had parental informed consent and child assent, however any participant aged between 5 and 13 years was included in statistical analysis in accordance with registered inclusion criteria.

Due to the effects of COVID-19, schools in the New York City area were closed to health services from March 2020 through September 2021. As a result, any schools enrolled prior to this period had follow-up care suspended until program operations were authorized to resume. Following this period, biannual screening and treatment resumed, and any newly enrolled schools were seen according to this schedule.

### Randomization

The unit of randomization was at the school level. Schools were block randomized to each treatment group using a random number generator.

### Data Collection

Prior to treatment, participants received a full visual-tactile oral examination at each study observation. Data were recorded for decay presence on every tooth surface (occlusal, lingual, buccal, mesial, and distal), missing at the tooth level, and filling presence. Dental caries was recorded if presenting as a score of 5 (distinct cavity with visible dentin) or 6 (extensive distinct cavity with visible dentin) on the International Caries Detection and Assessment System [14]. Additional data for socioeconomic indicators (e.g., age, sex, race/ethnicity) were obtained from informed consent documents. All data were stored locally on password-protected tablet computers until securely uploaded to a data coordinating center for processing and storage at the end of each study observation day.

### Interventions

All treatments provided in CariedAway were administered by either registered dental hygienists or registered nurses following didactic and experiential training with expert standardization. Children assigned to the experimental group received an application of 38% silver diamine fluoride (Elevate Oral Care Advantage Arrest 38%, 2.24 F-ion mg/dose) on posterior, asymptomatic, cavitated lesions and the pits and fissures of all sound bicuspids and molars. After cleaning and drying affected tooth surfaces, a microbrush was used to transfer SDF to individual teeth for a minimum of 30 seconds, followed by 60 seconds drying time. Subjects assigned to the active control received glass ionomer sealants (GC Fuji IX, GC America) on the pits and fissures of all sound bicuspids and molars, and the placement of atraumatic restorations on all frank, asymptomatic, cavitated lesions. Participants in both treatment groups then received fluoride varnish (5% NAF, Colgate PreviDent) applied to all teeth.

### Statistical Analysis

The analytic sample was restricted to participants who completed two or more post-baseline visits to ensure each child had a minimum of two observational periods for caries detection. No restrictions were placed on the time that elapsed between observations. Descriptive statistics were computed for the full and analytic samples for socioeconomic and clinical variables. Univariate failure time to first infection in the presence of interval-censored data was determined for the analytic sample using semiparametric transformation models [15], estimating the cumulative hazard function. Model parameters included treatment type, age at observation, the presence of untreated caries at baseline, and whether the subject had evidence of prior receipt of dental sealants at baseline. We then imputed caries occurrence conditional on these covariates using G-imputation [16] and estimated the per-visit caries incidence with 95% confidence intervals. Analysis was conducted in R v4.4.1. Statistical significance was determined at p < 0.05.

## Results

The full study sample consisted of 7418 participants enrolled in 48 schools (Table 1). There were 4006 (54.0%) females, and 66% were from either Black or Hispanic/Latino ethnicities. The average age at enrollment was 7.6 years (SD = 1.9), and the baseline prevalence of dental caries was 26.7% (95% CI = 25.7, 27.7). Following randomization, 2063 (50.3%) received silver diamine fluoride and 2037 (49.7%) received dental sealants and atraumatic restorations. For the analytic sample, there were 3040 participants remaining following exclusion criteria (1516 in the SDF group, 1524 in the sealant and ART group). There were 1687 males (55.5%), 1745 children from Hispanic/Latino race/ethnicities (57.4%), and 573 children from black race/ethnicities (18.9%). The average age for the analytic sample was 9.3 years (SD=1.8). A CONSORT participant enrollment flowchart is provided in Figure 1.

**Table 1:** Study enrollment and participant demographics.

|  | **Overall** |  | **SDF** |  | **Sealants** |  |
| --- | --- | --- | --- | --- | --- | --- |
|  | **N** | **%** | **N** | **%** | **N** | **%** |
| **Enrolled participants** | 7418 | 100 | 3739 | 50.4 | 3679 | 49.6 |
| Baseline caries | 1980 | 26.7 | 1016 | 27.2 | 964 | 26.2 |
| Sex (Males) | 3412 | 46 | 1785 | 47.7 | 1627 | 44.2 |
| Race/Ethnicity |  |  |  |  |  |  |
| Asian | 125 | 1.7 | 88 | 2.4 | 37 | 1 |
| Black | 1246 | 16.8 | 650 | 17.4 | 596 | 16.2 |
| Hispanic | 3648 | 49.2 | 1766 | 47.2 | 1882 | 51.2 |
| White | 153 | 2.1 | 86 | 2.3 | 67 | 1.8 |
| Multiple | 114 | 1.5 | 67 | 1.8 | 47 | 1.3 |
| Other | 90 | 1.2 | 56 | 1.5 | 34 | 0.9 |
| Unreported | 2042 | 27.5 | 1026 | 27.4 | 1016 | 27.6 |
| Age at baseline (mean/SD) | 7.6 | 1.9 | 7.5 | 1.9 | 7.6 | 1.9 |
| **Analyzed participants** | 3040 | 100 | 1516 | 49.9 | 1524 | 50.1 |
| Sex (Males) | 1351 | 44.44 | 812 | 53.56 | 647 | 42.45 |
| Race/Ethnicity |  |  |  |  |  |  |
| Asian | 53 | 1.74 | 42 | 2.77 | 11 | 0.72 |
| Black | 573 | 18.85 | 302 | 19.92 | 271 | 17.78 |
| Hispanic | 1745 | 57.4 | 859 | 56.66 | 886 | 58.14 |
| White | 69 | 2.27 | 46 | 3.03 | 23 | 1.51 |
| Multiple | 48 | 1.58 | 29 | 1.91 | 19 | 1.25 |
| Other | 41 | 1.35 | 22 | 1.45 | 19 | 1.25 |
| Unreported | 511 | 16.81 | 216 | 14.24 | 295 | 19.35 |
| Age at baseline (mean/SD) | 9.3 | 1.8 | 9.2 | 1.8 | 9.3 | 1.9 |

**Figure 1:**
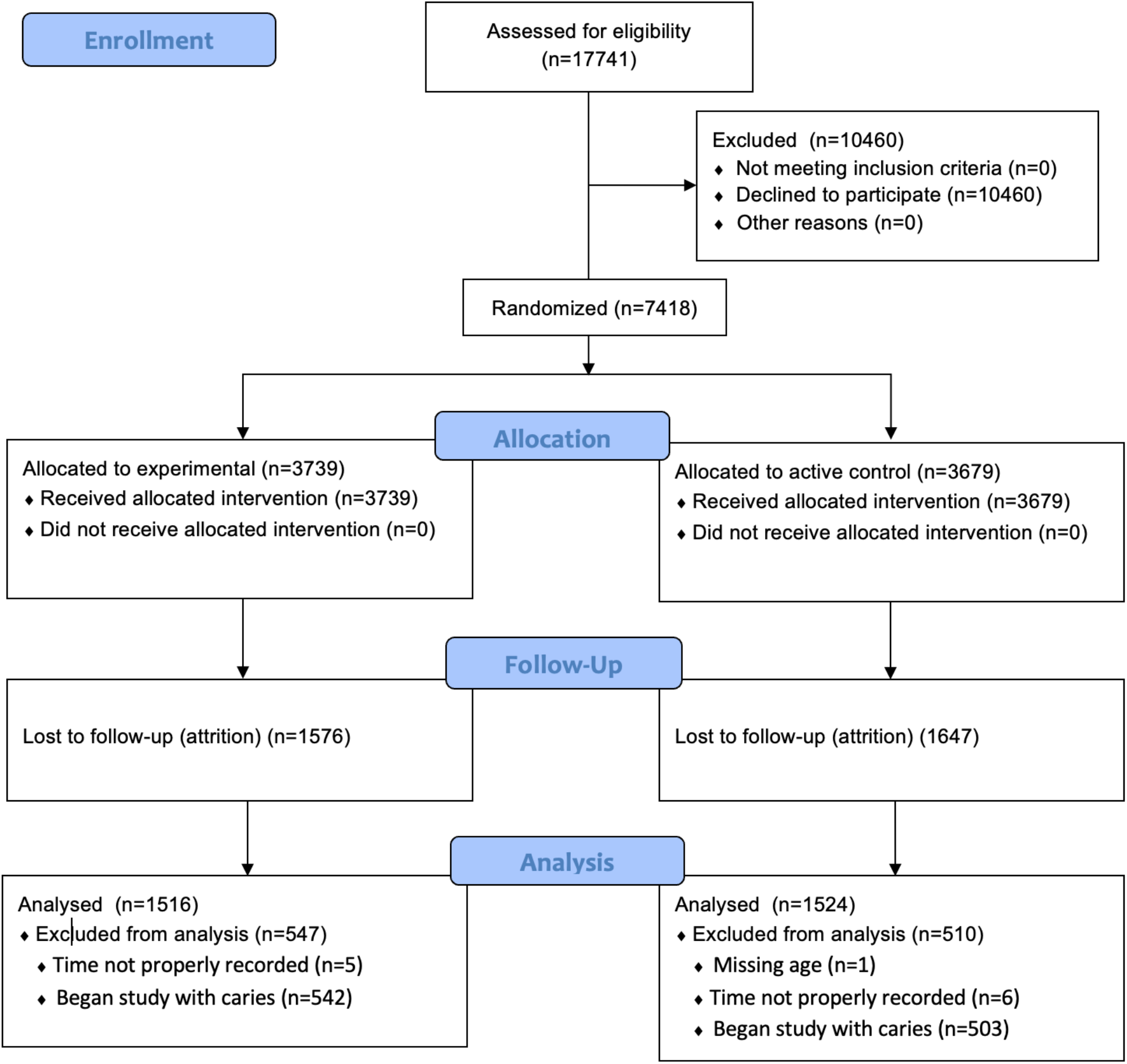
CONSORT Flowchart.

Results from adjusted failure models (Table 2) indicate no significant differences (p = 0.91) in the hazard of dental caries in children receiving SDF compared to dental sealants (HR = 0.99, 95% CI = 0.73, 1.24). Increases in age were associated with a slight decrease in the hazard of caries (HR = 0.97, 95% CI = -0.97, 0.98, p < .0001), and children with untread decay at baseline had a large increase in the hazard (HR = 2.54, 95% CI = 2.26, 2.83, p < .0001). There were no differences in the hazard of caries if children had any evidence of prior preventive care at baseline (p = 0.28). Following G-imputation (Table 3), the per-visit caries risk ranged from 4.79 per 1000-person years (95% CI = 4.13, 5.44) to 11.1 per 1000-person years (95% CI = 5.82, 16.37) post-baseline, with the incidence increasing with each successive observation.

**Table 2:** Hazard of dental caries by select covariates (univariate failure time)

|  | HR | 95% L HR | 95% U HR | Z-Stat | P-value |
| --- | --- | --- | --- | --- | --- |
| Treatment (SDF vs sealants) | 0.9853307 | 0.7269221 | 1.2437394 | -0.112088 | 0.9107531 |
| Age | 0.9743498 | 0.9657073 | 0.9829924 | -5.892965 | < .0001 |
| Baseline decay | 2.5413837 | 2.2555742 | 2.8271932 | 6.396251 | < .0001 |
| Baseline sealants | 1.2949162 | 0.8282655 | 1.7615670 | 1.085510 | 0.2776957 |

**Table 3:** Caries incidence per 1000-person years (by visit)

| Follow-up | Incidence | SE | 95% L | 95% U |
| --- | --- | --- | --- | --- |
| 1st | 6.4495E-07 | 2.4953E-04 | -4.8843E-04 | 4.8972E-04 |
| 2nd | 4.7870E+00 | 3.3532E-01 | 4.1298E+00 | 5.4442E+00 |
| 3rd | 7.1998E+00 | 1.6518E+00 | 3.9622E+00 | 1.0437E+01 |
| 4th | 1.1094E+01 | 2.6911E+00 | 5.8201E+00 | 1.6369E+01 |

## Discussion

In this pragmatic clinical trial of minimally invasive interventions for dental caries provided in a school-based model, we found no significant differences in the hazard of caries when comparing children treated with either silver diamine fluoride and fluoride varnish to glass ionomer sealants and fluoride varnish. Additionally, the per-visit caries incidence increased with each successive observation and was significantly different from zero, but the overall incidence was low.

Data on caries incidence in United States children participating in school dental health programs is lacking in the epidemiologic literature, although caries experience (treated and untreated caries) in children aged 6-11 years was previously reported to be 50.5% using data from the 2015-2016 National Health and Nutrition Examination Survey [17]. In other studies, the incidence of the first carious lesion in primary molars was 9.5 per 100 person-years in four-year old English children, with caries incidence in those with existing caries at recruitment being five times greater than those without [18], and incidence in the first primary molars in Iranian children aged 7-8 years was 0.16 [19]. Similarly, data from US preschool children participating in a school readiness program for low-income families showed that the two-year incidence of dental caries (measured as decayed, missing, and filled surfaces) was 1.4 in caries-free children, compared to 2.9 in children with pit/fissure caries at baseline and 5.1 in children with maxillary anterior caries at baseline [20]. Our results support similar conclusions, in that children with preexisting caries had significantly higher risk of further incidence. Overall, the 2019 incidence rate of caries in permanent teeth in children aged 5-14 years was 30.6% in the high-income North America region, compared to 31.6% in 1990, indicating only small decreases over thirty years [21].

The World Health Organization considers schools to be an important factor in addressing child and adolescent health, publishing a “Health Promoting Schools Framework” for healthy behaviors and engaging with families and communities. Schools are promising environments for health promotion whether initiated by local school practices or top-down policies [22], but careful implementation research is necessary to reach their potential [23], especially if the goal is to reduce health disparities. The typical typology of health interventions that are implemented in schools include health promotion policies, environmental change strategies, health education messaging, and delivery of prevention and clinical care. For this latter type, established programs include immunization, vision, mental health assessment, and contraception. However, there is a clear need for sustainable school health programs focused on oral disease prevention. Both the 2000 and 2020 U.S. Department of Health and Human Services reports on the oral health of America highlighted the need for and efficacy of caries prevention. Between the publication of these reports, the National Academy of Medicine (then known as the Institute of Medicine) identified the comparative effectiveness of caries prevention programs as a “high priority” topic [24], and the CDC published the *Oral Health In America: Summary of the Surgeon General’s Report*, concluding that oral health is integral to the overall health and well-being of all Americans.

There are a number of limitations for the current study. Although data were derived from a randomized clinical trial, there was considerable attrition and a number of study participants were not seen for a follow-up observation, likely due to the disruptions in program operations due to COVID-19. Furthermore, this suspension of care meant that annual caries incidence could not be computed. However, our reporting of the per-visit incidence may be equally useful in determining the expected impact of a school caries prevention program. Finally, our focus on the incidence of caries on any dentition, as opposed to first primary molars only or other tooth-specific stratification, reflects a public health approach to community caries prevention. This likely inflates the caries incidence and thus may limit the extrapolation of results to other contexts. Future additions to this research can restrict analysis to first or second molars, and also consider multiple failure events (e.g., multiple teeth failing or failures over multiple observational periods), which can also be accounted in the methods used in the present study [15] .

Prior evidence from school-based caries prevention suggests that both dental sealants/ART [11, 25] and SDF [10] can prevent and control dental caries. However, much of the established literature did not provide for any longitudinal follow-up of program participants, nor did any estimates of program impact account for the interval censoring in caries incidence. We used recent methods for interval censored data and multiple imputation for disease incidence, while including data that spanned a five-year caries prevention program operating in high-risk elementary schools in an urban district. Our results can inform future program planning and implementation for school oral health programs, as the incidence rate of caries shows a progressive increase in time following initialization.

## Data Availability

All data produced in the present study are available upon reasonable request to the authors after an approved proposal and data use agreemtent, following the schedule of data availability previously published for this trial.

